# Association of preserved ratio impaired spirometry with mortality and airflow obstruction in the silicotics: a longitudinal cohort study

**DOI:** 10.1101/2024.06.27.24309566

**Authors:** Shuyuan Yang, Lap Ah Tse

**Affiliations:** Jockey Club School of Public Health and Primary Care, The Chinese University of Hong Kong, Hong Kong SAR

**Keywords:** Lung function, preserved ratio impaired spirometry, airflow obstruction, mortality, silicosis

## Abstract

**Rationale:** Preserved ratio impaired spirometry (PRISm), defined as an impaired forced expiratory volume in one second (FEV_1_) with a preserved ratio of FEV_1_ to forced vital capacity (FVC), is associated with increased risk of airflow obstruction (AFO) and mortality in the general population. However, evidence is limited among the individuals with silicosis, an old occupational disease with an ongoing outbreak in some developed countries.

**Objectives:** To investigate the association of PRISm with the risk of mortality and incident AFO in a cohort of workers with silicosis.

**Methods:** A total of 4315 workers aged 18-80 years and diagnosed with silicosis at the Pneumoconiosis Clinic, Tuberculosis and Chest Service during 1981-2019 were enrolled in this study and followed up for a median of 12.3 years till 31 December 2019. Spirometry was included in the diagnostic examination of silicosis and follow-up reassessments. Lung function categories of participants were classified as normal spirometry (FEV_1_/FVC ≥ 0.7, FEV_1_ ≥ 80% predicted), PRISm (FEV_1_/FVC ≥ 0.7, FEV_1_ < 80% predicted), and AFO (FEV_1_/FVC < 0.7). The hazard ratio (HR) and 95% confidence intervals (95% CI) were estimated using Cox proportional hazards models adjusting for age, body mass index, tuberculosis history, smoking, and radiographic characteristics.

**Measurements and Main Results:** During the follow-up period, a total of 2399 (55.6%) subjects died, 1359 of whom died from respiratory-related diseases, and 780 subjects developed AFO. Subjects with PRISm had significantly increased multivariable-adjusted risk of all-cause death (adjusted HR=1.63, 95% CI 1.44-1.85) and respiratory-related mortality (adjusted HR=1.74, 95% CI 1.48-2.05) as compared with the those with normal spirometry. Besides, there was a higher risk of developing AFO in subjects with PRISm than in those with normal spirometry (adjusted HR=1.46, 95% CI 1.22-1.75). No significant interaction was observed between PRISm and smoking status in the risk of all-cause mortality and incident AFO.

**Conclusions:** PRISm is significantly associated with increased all-cause and respiratory-related mortality and a greater risk of progression to AFO among the individuals with silicosis.

## Introduction

Chronic obstructive pulmonary disease (COPD) is among the most important contributors to the increasing burden of non-communicable diseases worldwide^1,2^. According to the Global Burden of Disease Study 2019, the prevalent cases of COPD reached 212.3 million, while COPD accounted for 3.3 million deaths and 74.4 million disability adjusted life years around the world^3^. COPD is characterized by airflow obstruction (AFO) that defined as an impaired ratio of forced expiratory volume in one second (FEV_1_) to forced vital capacity (FVC) less than 0.7 or lower limit of normal on spirometry^4,5^. However, preserved ratio impaired spirometry (PRISm), a non-obstructive ventilatory defect that characterized by simultaneously deficits in both FEV_1_ and FVC but with preserved ratio of FEV_1_/FVC, was often neglected in the clinical setting in the past but now attracted a growing attention^6^. Recent epidemiologic studies in the general population revealed that PRISm is associated with both impaired quality of life and poor clinical outcomes, e.g., all-cause death, respiratory-related events (e.g., hospitalizations and mortality), and cardiovascular-related events^7–10^. Although the biological mechanism of PRISm and its clinical prognosis are poorly understood, the recognition of this lung function category is critical for physicians in developing clinical management strategies. Despite the considerable prevalence rate of PRISm from 7.1% to 25.9% globally^11,12^, guidelines for the diagnostic evaluation and clinical management of PRISm have not yet been well established due to the lack of pertinent evidence for policy making^10,13,14^.

Silicosis is one of the most important occupational diseases worldwide caused by the prolonged inhalation of respirable silica dust. Despite the tremendous efforts for decades in minimizing the occurrence of silicosis, failure to recognize and eliminate the silica-related exposure in some contemporary work practices, e.g., denim jean production, domestic benchtop fabrication and jewelry polishing, leaded to a global re-emergence of this ancient and potentially fatal pneumoconiosis in some developed countries ^15,16^. Although PRISm has been associated with long-term adverse clinical outcomes in previous general population-based studies^9,10,17–21^, evidence is lacking among the silicotics with low compliance of lung caused by the replacement of elastin fibers by collagens in the nodular fibrosis^22^. To address this knowledge gap, the present study aimed to investigate the association between PRISm and the risk of all-cause and cause-specific mortality in the silicotics and estimate the risk of developing AFO among the silicotics with PRISm using the data from a large occupational cohort in Hong Kong from 1981 to 2019.

## Methods

### Study population and data

This is a retrospective cohort study using the data from a territory-wide silicosis cohort of consecutive workers who were diagnosed with silicosis in Hong Kong during 1981-2019. The details of this cohort have been described elsewhere^23–26^. Briefly, this cohort contains 4481 workers who were diagnosed with silicosis at the Pneumoconiosis Clinic, Tuberculosis and Chest Service from 1981 to 2019. The median time between the diagnostic examination of silicosis (baseline spirometry) and death or the study end day (i.e., December 31, 2019) was 12.3 years (interquartile range: 5.9 to 19.4 years). Silicosis was diagnosed based on the radiographic signs of nodular fibrosis (silicotic nodules) in combination with a work history involving occupational exposure to silica-related dusts ^27^. The radiographic changes were determined independently by three members of the Pneumoconiosis Medical Board according to the presence of round and/or irregular small opacities with diameters up to 1.5 mm, 1.5-3 mm, or 3-10 mm (i.e., p, q, and r (for round opacities)/s, t, and u (for irregular opacities) according to the International Labor Office Classification) or large opacities having the longest dimension up to 50 mm (progressive massive fibrosis)^28^. Once diagnosed with silicosis, the workers were entitled to the pneumoconiosis compensation assessments containing extensive clinical interviews concerning symptoms, lifestyle, health topics and work history, physical health examinations including spirometry will also be performed hereafter. Individuals with at least one valid spirometry were retained for analyses, and the first spirometry obtained was considered as baseline spirometry. This study was approved by the Survey and Behavior Research Ethics Committee of The Chinese University of Hong Kong (SBRE-19-023).

### Spirometry

The parameters of lung function, including forced expiratory volume in one second (FEV_1_) and forced vital capacity (FVC), were measured using a wedge-type bellow spirometer (Vitalograph PFT II plus, Buckingham, UK) following the American Thoracic Society/European Respiratory Society guidelines. The nurses took three readings of FEV_1_ and FVC corrected for body temperature, water vapor saturation and pressure from satisfactory maneuvers, and only the best one was recorded for analyses. We calculated the predicted values of FEV_1_ and FVC and lower limit of normal (LLN) of FEV_1_/FVC using the reference equations published by the Hong Kong Thoracic Society for the local population^29,30^. The lung function categories were defined according to the Global Initiative for Chronic Obstructive Lung Disease (GOLD) criteria and previous reports^10–12,31–34^ as follows: (1) normal spirometry: FEV_1_/FVC ≥ 0.70, FEV_1_ ≥ 80% predicted; (2) PRISm: FEV_1_/FVC ≥ 0.70, FEV_1_ ≤ 80% predicted; (3) AFO GOLD 1: FEV_1_/FVC < 0.70, FEV_1_ ≥ 80% predicted; (4) AFO GOLD 2: FEV_1_/FVC < 0.70, 50% ≤ FEV_1_ < 80% predicted; (5) AFO GOLD 3: FEV_1_/FVC < 0.70, 30% ≤ FEV_1_ < 50% predicted; (6) AFO GOLD 4: FEV_1_/FVC < 0.70, FEV_1_ ≤ 30% predicted.

### Outcomes

The main outcomes were (1) all-cause mortality (ICD-10: A00-Z99); (2) respiratory-related mortality (ICD-10: J00-J99); (3) lung cancer mortality (ICD-10: C34); (4) cardiovascular-related mortality (ICD-10: I00-I99); and (5) incident AFO. When a subject died, all medical information relating to the death, e.g., the date and underlying cause of death, were extracted from the medical records, while the official death certificate issued by a registered medical partitioner who attended this subject’s last illness was also extracted. We coded underlying and contributing causes of death according to the International Statistical Classification of Diseases and Related Health Problem, 10^th^ Revision (ICD-10). The development of AFO was determined by the reassessment of spirometry during the follow-up period.

### Statistical analyses

We estimated the silica dust exposure for each episode of job by multiplying the exposure level of the certain job by job duration after linking the individual occupation to the job-exposure matrix that developed based on the exposure levels summarized by the Occupational Safety and Health Administration (OSHA)^35^. The cumulative dust exposure (mg/m^3^-year) was then obtained by summing up the estimated exposure of all episodes of jobs, as in equation (1).

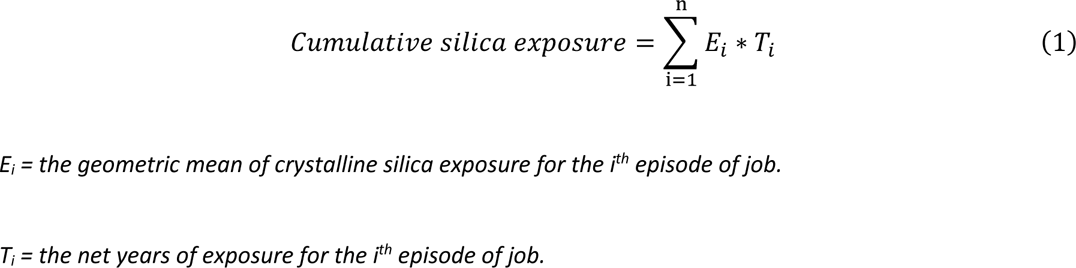

Univariate comparisons between subjects with different lung function categories were made using Chi-square tests. The cumulative survival rates for all-cause death were calculated using Kaplan-Meier method and compared using a log-rank test. The hazard ratios and their 95% confidence intervals for all-cause death, respiratory-related death, lung cancer death, cardiovascular-related death and incident AFO were estimated using Cox proportional hazard regression or Fine-Gray competing risk model^36^ (a natural extension of the Cox regression to the settings of cause-specific mortality with competing risks) that adjusting for age, body mass index, history of tuberculosis, cumulative silica exposure, smoking status, pack-years, and radiographic signs of silicotic nodules. We also conducted subgroup analyses by the shape, size, or profusion of small opacities and the presence of progressive massive fibrosis. To ensure the robustness of results, the key analyses were carried out based on LLN-defined PRISm (FEV_1_/FVC ≥ LLN, FEV_1_ ≤ 80% predicted). SAS software package version 9.4 (SAS Institute, Cary, North Carolina) was used to carry out all statistical analyses. A two-side *p* < 0.05 was considered statistically significant.

## Results

### Baseline prevalence and characteristics of PRISm

Among 4481 subjects who diagnosed with silicosis at the Pneumoconiosis Clinic during 1981-2019, 166 (3.7%) were excluded because they were aged over 80 years old (due to the limitation of prediction formulae for reference values), without baseline physical examination or invalid spirometry results **(Table E1)**. The prevalence of PRISm among the remaining 4315 subjects with valid baseline spirometry was 11.8% (n=508). Characteristics of the subjects with PRISm relative to those with normal spirometry and AFO are shown in **Table 1**. Subjects with PRISm had more average pack-years and respiratory symptoms, e.g., dyspnea and hemoptysis, as compared with those with normal spirometry.

**Table 1.**
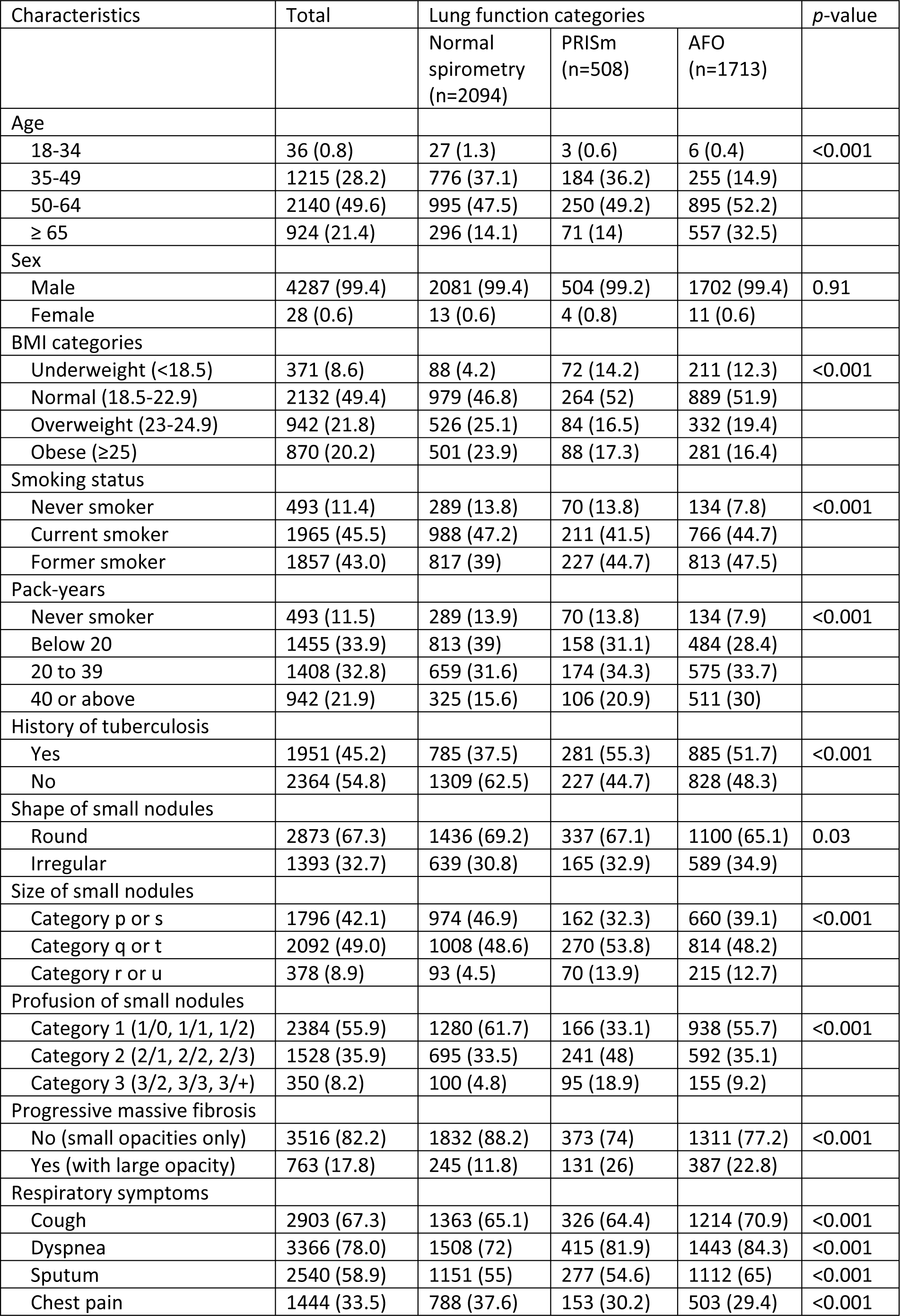
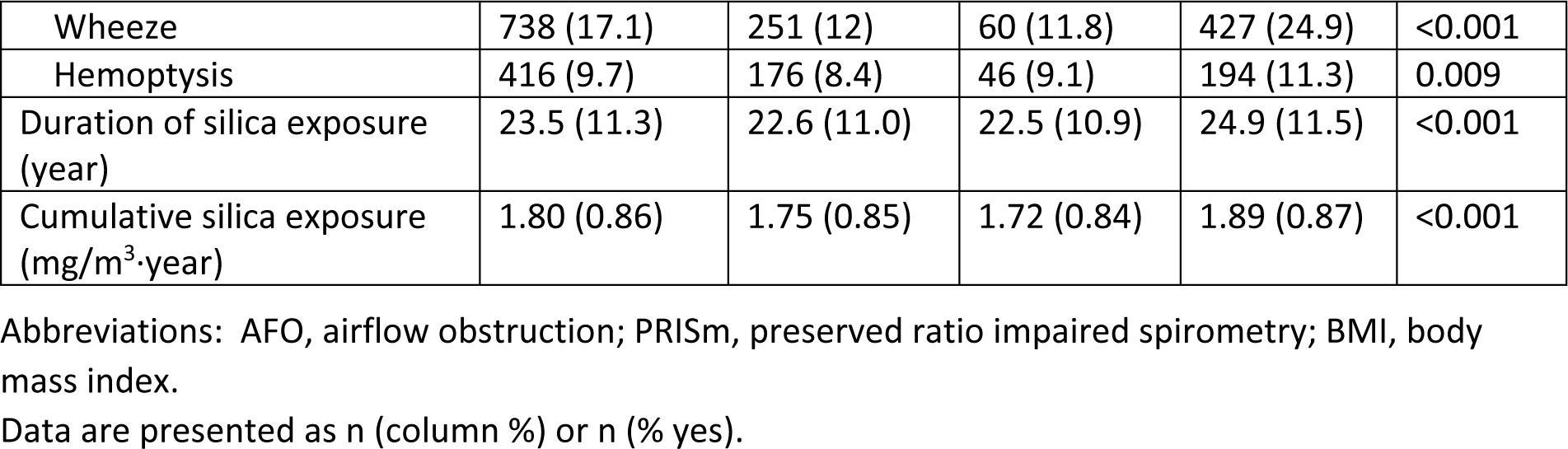
Baseline characteristics of the cohort (n=4315) by spirometric lung function class.

### Mortality by lung function category

Subjects with valid baseline lung function data and subsequent assessment of vital status available (n=4315) were included in the analysis of mortality rates by lung function category, and 2399 (55.6%) subjects among them died during the follow-up period. The crude cumulative survival rate in subjects with PRISm or AFO was significantly lower than those with normal spirometry, as shown in **Figure 1**. Hazard ratios (HR) with 95% confidence interval (CI) from Cox proportional hazard models are shown in **Table 2**. The risk of all-cause death was significantly higher in subjects with PRISm than in those with normal spirometry (crude HR=1.84, 95% CI: 1.63-2.08), and PRISm remained a significant predictor after adjusting for age, BMI, tuberculosis, cumulative silica exposure, smoking habits, and radiographic signs (adjusted HR=1.63, 95% CI: 1.44-1.85). Subjects with restrictive PRISm had an adjusted risk of all-cause death (adjusted HR=1.76, 95% CI: 1.53-2.02) intermediate between those with GOLD2 and GOLD 3 spirometry, which was higher than those with non-restrictive PRISm (adjusted HR=1.31, 95% CI: 1.03-1.66) **(Table E2)**. For cause-specific death, we found significantly greater risk of respiratory-related mortality among subjects with PRISm or AFO (adjusted HR=1.74, 95% CI: 1.48-2.05 for PRISm), but no significant association between PRISm and mortality from lung cancer or cardiovascular diseases was observed. In a subgroup analysis of subjects by shape, profusion, and size of silicotic opacities as well as progressive massive fibrosis, those with PRISm had a significant increased risk of all-cause mortality among all subgroups, as shown in **Figure 2** and **Table E3-6**. As shown in **Table E7**, LLN-defined PRISm was also significantly associated with increased risk of all-cause and respiratory-related mortality, which was consistent with the association between mortality and PRISm based on the fixed cut-off.

**Table 2.**
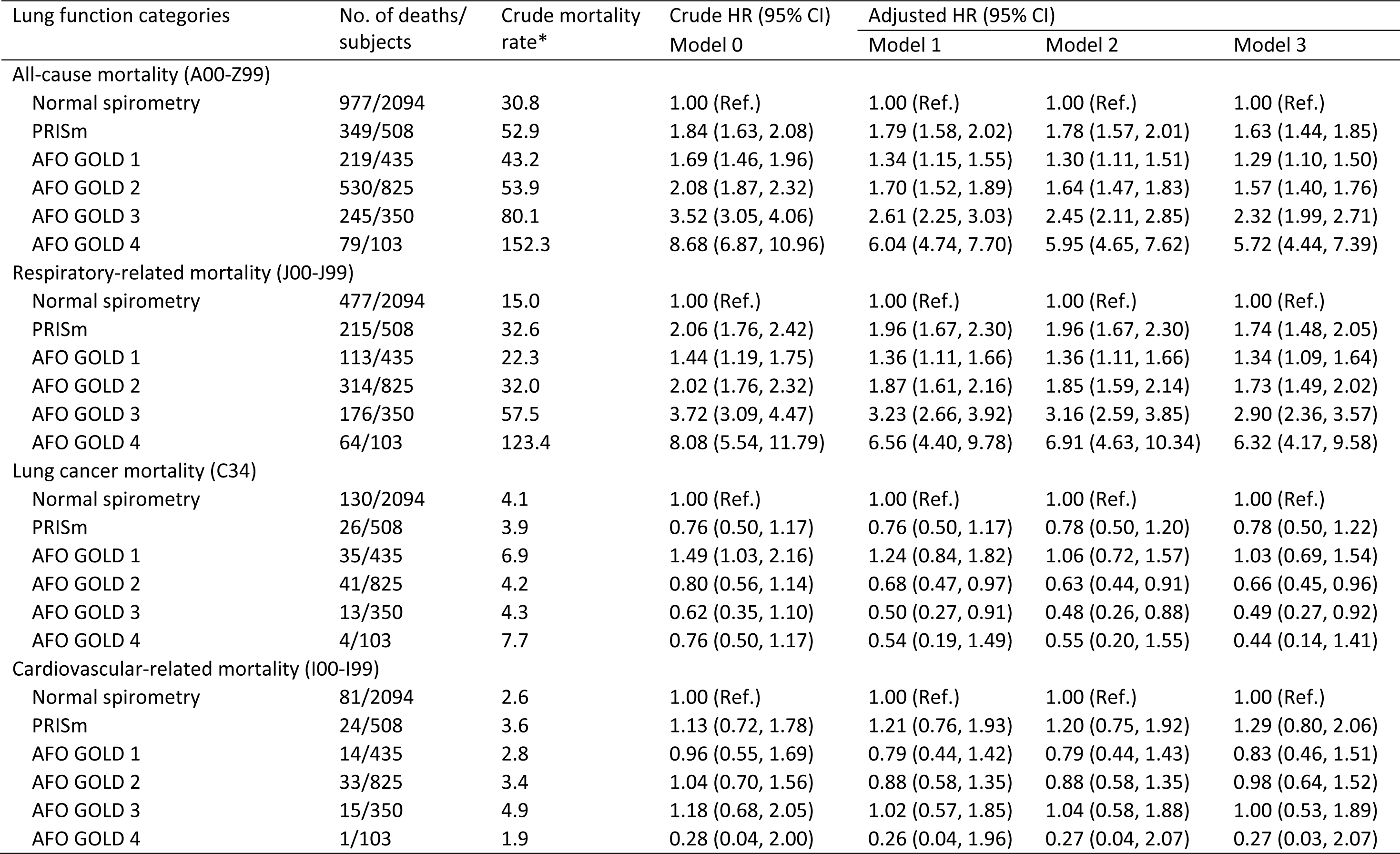
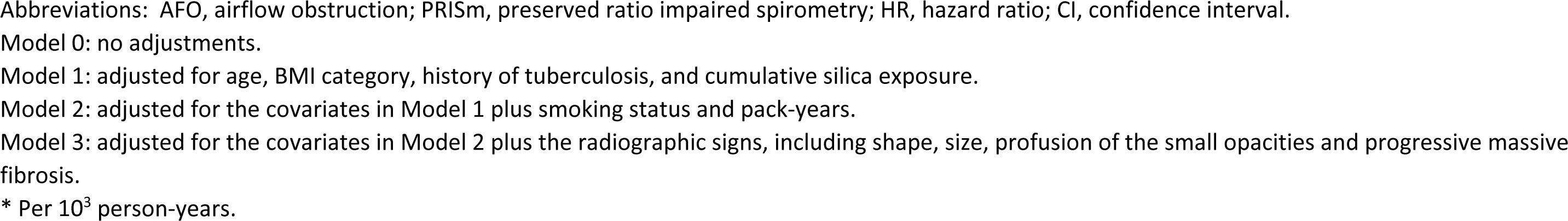
Hazard ratios (HR) and 95% confidence intervals (CI) for all-cause and major cause-specific mortality by spirometric lung function class.

| Lung function categories | No. of deaths/<br>subjects | Crude mortality<br>rate* | Crude HR (95% CI)<br>Model 0 | Adjusted HR (95% CI) |  |  |
| --- | --- | --- | --- | --- | --- | --- |
|  |  |  |  | Model 1 | Model 2 | Model 3 |
| All-cause mortality (A00-Z99) |  |  |  |  |  |  |
| Normal spirometry | 977/2094 | 30.8 | 1.00 (Ref.) | 1.00 (Ref.) | 1.00 (Ref.) | 1.00 (Ref.) |
| PRISm | 349/508 | 52.9 | 1.84 (1.63, 2.08) | 1.79 (1.58, 2.02) | 1.78 (1.57, 2.01) | 1.63 (1.44, 1.85) |
| AFO GOLD 1 | 219/435 | 43.2 | 1.69 (1.46, 1.96) | 1.34 (1.15, 1.55) | 1.30 (1.11, 1.51) | 1.29 (1.10, 1.50) |
| AFO GOLD 2 | 530/825 | 53.9 | 2.08 (1.87, 2.32) | 1.70 (1.52, 1.89) | 1.64 (1.47, 1.83) | 1.57 (1.40, 1.76) |
| AFO GOLD 3 | 245/350 | 80.1 | 3.52 (3.05, 4.06) | 2.61 (2.25, 3.03) | 2.45 (2.11, 2.85) | 2.32 (1.99, 2.71) |
| AFO GOLD 4 | 79/103 | 152.3 | 8.68 (6.87, 10.96) | 6.04 (4.74, 7.70) | 5.95 (4.65, 7.62) | 5.72 (4.44, 7.39) |
| Respiratory-related mortality (J00-J99) |  |  |  |  |  |  |
| Normal spirometry | 477/2094 | 15.0 | 1.00 (Ref.) | 1.00 (Ref.) | 1.00 (Ref.) | 1.00 (Ref.) |
| PRISm | 215/508 | 32.6 | 2.06 (1.76, 2.42) | 1.96 (1.67, 2.30) | 1.96 (1.67, 2.30) | 1.74 (1.48, 2.05) |
| AFO GOLD 1 | 113/435 | 22.3 | 1.44 (1.19, 1.75) | 1.36 (1.11, 1.66) | 1.36 (1.11, 1.66) | 1.34 (1.09, 1.64) |
| AFO GOLD 2 | 314/825 | 32.0 | 2.02 (1.76, 2.32) | 1.87 (1.61, 2.16) | 1.85 (1.59, 2.14) | 1.73 (1.49, 2.02) |
| AFO GOLD 3 | 176/350 | 57.5 | 3.72 (3.09, 4.47) | 3.23 (2.66, 3.92) | 3.16 (2.59, 3.85) | 2.90 (2.36, 3.57) |
| AFO GOLD 4 | 64/103 | 123.4 | 8.08 (5.54, 11.79) | 6.56 (4.40, 9.78) | 6.91 (4.63, 10.34) | 6.32 (4.17, 9.58) |
| Lung cancer mortality (C34) |  |  |  |  |  |  |
| Normal spirometry | 130/2094 | 4.1 | 1.00 (Ref.) | 1.00 (Ref.) | 1.00 (Ref.) | 1.00 (Ref.) |
| PRISm | 26/508 | 3.9 | 0.76 (0.50, 1.17) | 0.76 (0.50, 1.17) | 0.78 (0.50, 1.20) | 0.78 (0.50, 1.22) |
| AFO GOLD 1 | 35/435 | 6.9 | 1.49 (1.03, 2.16) | 1.24 (0.84, 1.82) | 1.06 (0.72, 1.57) | 1.03 (0.69, 1.54) |
| AFO GOLD 2 | 41/825 | 4.2 | 0.80 (0.56, 1.14) | 0.68 (0.47, 0.97) | 0.63 (0.44, 0.91) | 0.66 (0.45, 0.96) |
| AFO GOLD 3 | 13/350 | 4.3 | 0.62 (0.35, 1.10) | 0.50 (0.27, 0.91) | 0.48 (0.26, 0.88) | 0.49 (0.27, 0.92) |
| AFO GOLD 4 | 4/103 | 7.7 | 0.76 (0.50, 1.17) | 0.54 (0.19, 1.49) | 0.55 (0.20, 1.55) | 0.44 (0.14, 1.41) |
| Cardiovascular-related mortality (I00-I99) |  |  |  |  |  |  |
| Normal spirometry | 81/2094 | 2.6 | 1.00 (Ref.) | 1.00 (Ref.) | 1.00 (Ref.) | 1.00 (Ref.) |
| PRISm | 24/508 | 3.6 | 1.13 (0.72, 1.78) | 1.21 (0.76, 1.93) | 1.20 (0.75, 1.92) | 1.29 (0.80, 2.06) |
| AFO GOLD 1 | 14/435 | 2.8 | 0.96 (0.55, 1.69) | 0.79 (0.44, 1.42) | 0.79 (0.44, 1.43) | 0.83 (0.46, 1.51) |
| AFO GOLD 2 | 33/825 | 3.4 | 1.04 (0.70, 1.56) | 0.88 (0.58, 1.35) | 0.88 (0.58, 1.35) | 0.98 (0.64, 1.52) |
| AFO GOLD 3 | 15/350 | 4.9 | 1.18 (0.68, 2.05) | 1.02 (0.57, 1.85) | 1.04 (0.58, 1.88) | 1.00 (0.53, 1.89) |
| AFO GOLD 4 | 1/103 | 1.9 | 0.28 (0.04, 2.00) | 0.26 (0.04, 1.96) | 0.27 (0.04, 2.07) | 0.27 (0.03, 2.07) |

Abbreviations: AFO, airflow obstruction; PRISm, preserved ratio impaired spirometry; HR, hazard ratio; CI, confidence interval.
Model 0: no adjustments.
Model 1: adjusted for age, BMI category, history of tuberculosis, and cumulative silica exposure.
Model 2: adjusted for the covariates in Model 1 plus smoking status and pack-years.
Model 3: adjusted for the covariates in Model 2 plus the radiographic signs, including shape, size, profusion of the small opacities and progressive massive fibrosis.
\* Per 10<sup>3</sup> person-years.

**Figure 1.**
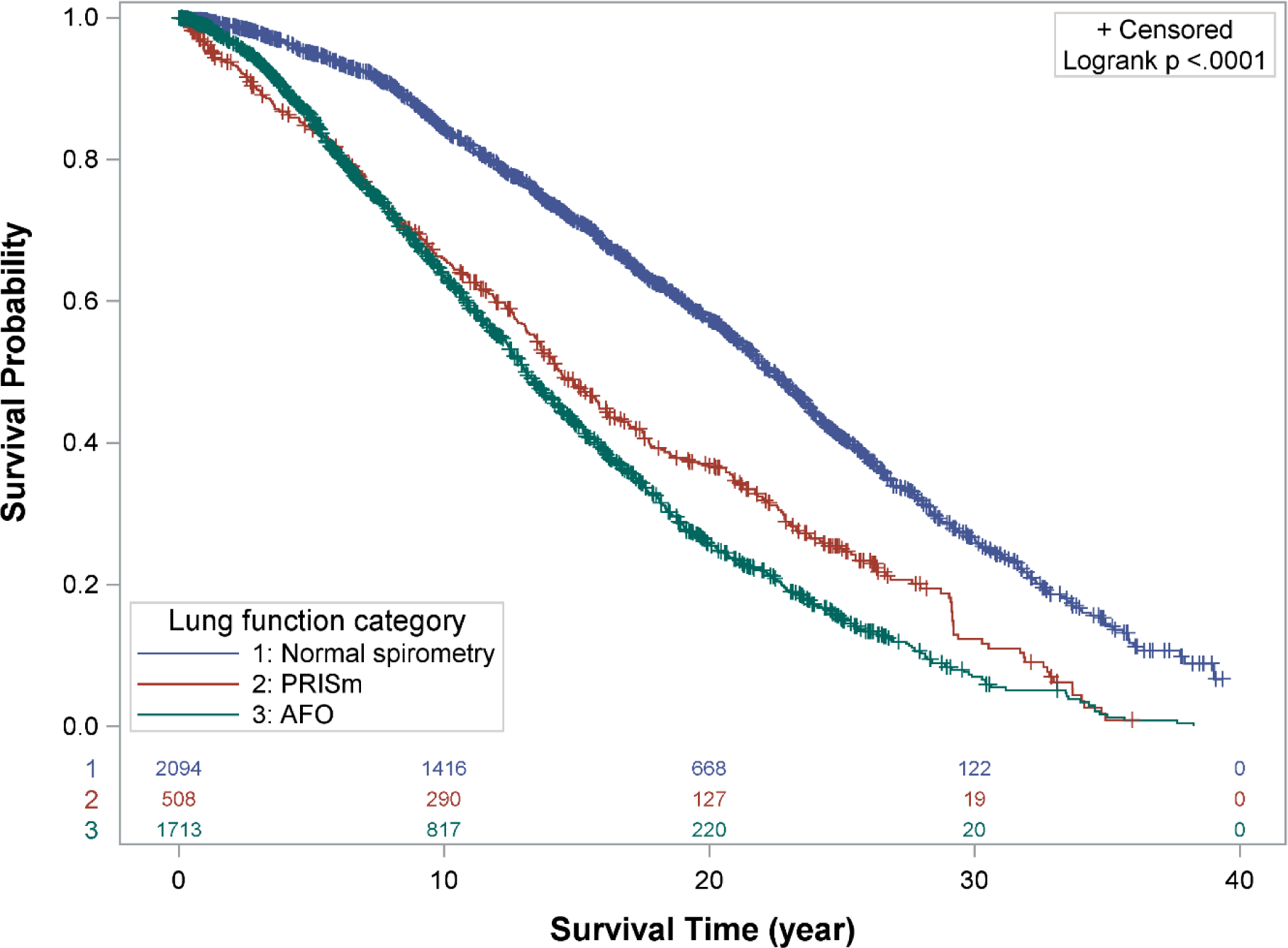
Kaplan-Meier curves of all-cause mortality with 95% confidence interval (CI) by spirometric lung function class Abbreviations: AFO, airflow obstruction; PRISm, preserved ratio impaired spirometry.

**Figure 2.**
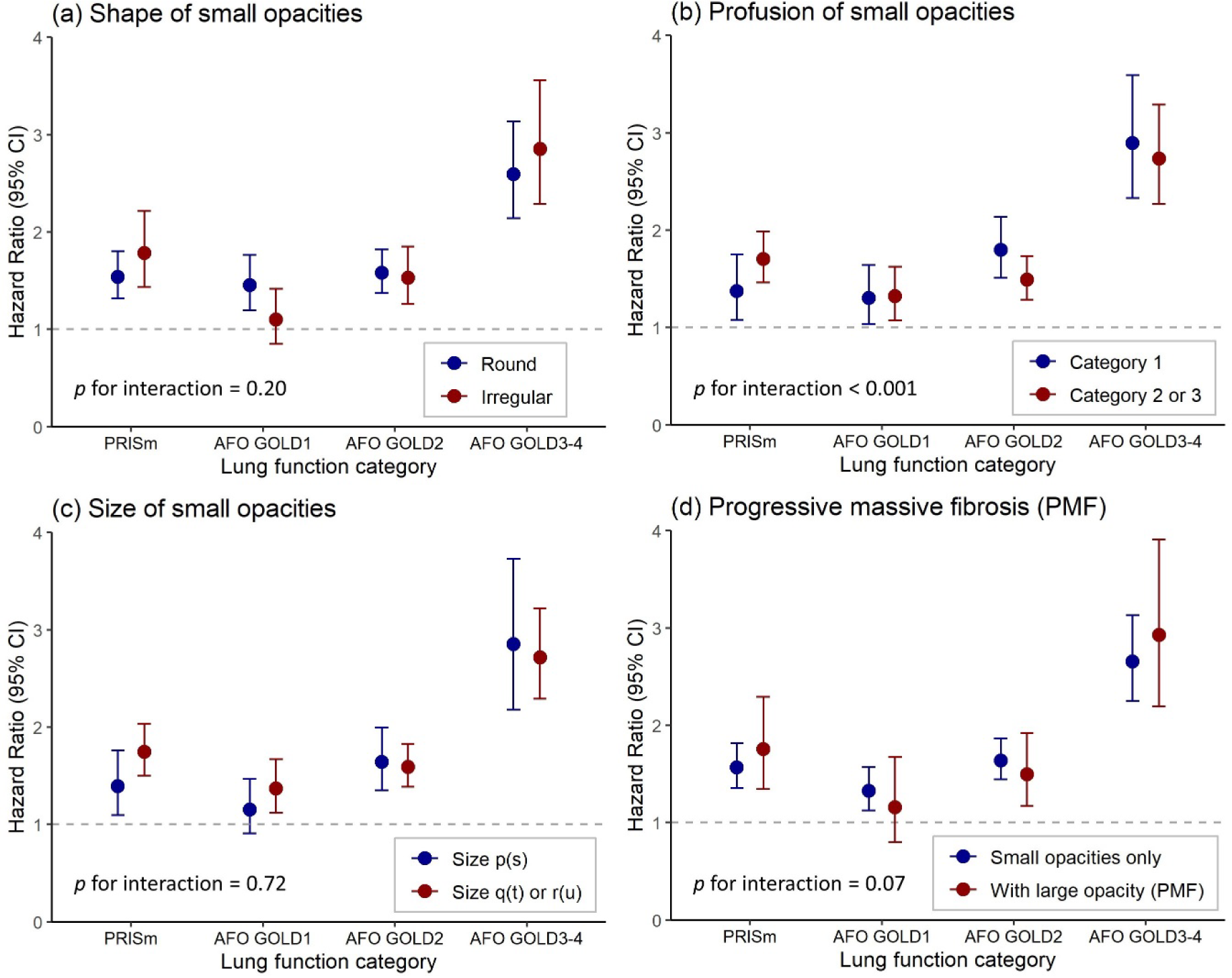
Association of preserved ratio impaired spirometry (PRISm) and airflow obstruction (AFO) with all-cause mortality by (a) shape, (b) profusion, and (c) size of silicotic opacities and (d) progressive massive fibrosis All hazard ratios were adjusted for age, BMI, smoking status, pack-years, tuberculosis, and cumulative silica exposure. Subjects with normal spirometry were used as reference. Abbreviations: AFO, airflow obstruction; PRISm, preserved ratio impaired spirometry; PMF, progressive massive fibrosis.

**Table 3.** Hazard ratios (HR) and 95% confidence intervals (CI) for the development of airflow obstruction.

| Lung function categories | No. of events/<br>subjects | Crude HR (95% CI) | Adjusted HR (95% CI) |  |  |
| --- | --- | --- | --- | --- | --- |
|  |  | Model 0 | Model 1 | Model 2 | Model 3 |
| Normal spirometry | 607/1452 | 1.00 (Ref.) | 1.00 (Ref.) | 1.00 (Ref.) | 1.00 (Ref.) |
| PRISm | 173/331 | 1.64 (1.39, 1.95) | 1.58 (1.33, 1.87) | 1.56 (1.31, 1.86) | 1.46 (1.22, 1.75) |

Abbreviations: PRISm, preserved ratio impaired spirometry; HR, hazard ratio; CI, confidence interval.
Model 0: no adjustments.
Model 1: adjusted for age, BMI category, history of tuberculosis, and cumulative silica exposure.
Model 2: adjusted for the covariates in Model 1 plus smoking status and pack-years.
Model 3: adjusted for the covariates in Model 2 plus the radiographic signs, including shape, size, profusion of the small opacities and progressive massive fibrosis.
\* Per 10<sup>3</sup> person-years.

### Cumulative incidence of AFO

The risk of developing AFO in PRISm was estimated in 1783 subjects without AFO at baseline who underwent at least one follow-up spirometry, among whom 41.8% (n=607) of the subjects with normal spirometry and 52.2% (n=173) of the subjects with PRISm developed AFO during the study period. Subjects with PRISm had significantly increased risk of developing AFO (crude HR=1.64, 95% CI: 1.39-1.95) as compared to those with normal spirometry at baseline. This association remained significant after adjusting for age, BMI category, tuberculosis history and cumulative silica exposure (model 1, adjusted HR=1.58, 95% CI: 1.33-1.87), for smoking status and pack-years in addition to covariates in model 1 (model 2, adjusted HR=1.56, 95% CI: 1.31-1.86), and for radiographic signs of silicotic nodules, e.g., shape, size and profusion of small opacities and presence of progressive massive fibrosis, other than those in model 2 (model 3, adjusted HR=1.46, 95% CI: 1.22-1.75). The robust association between PRISm and increased risk of incident AFO was observed in the sensitivity analysis using LLN threshold of FEV1/FVC ratio as the criterion for AFO (data not shown). The annual rate of decline in FEV1/FVC ratio was significantly greater in subjects with PRISm than that in subjects with normal spirometry (-0.0095/year for normal spirometry; -0.0138/year for PRISm; p=0.003 by Wilcoxon test).

### Combined influence of PRISm and smoking on risk of all-cause death and incident AFO

The combined influence of PRISm and smoking status on the risk of all-cause mortality and the development of AFO is shown **Figure 3**. Compared with the reference group of non-smokers with normal spirometry, subjects with PRISm had a significantly increased risk of all-cause mortality no matter what smoking status was, with a decreased risk among smokers who had quitted the habit of smoking (adjusted HR=1.79, 95% CI: 1.12-1.85 for never smokers with PRISm; adjusted HR=1.84, 95% CI:1.33-2.54 for former smokers with PRISm; adjusted HR=2.25, 95% CI:1.63-3.10 for current smokers with PRISm). The risk of AFO was also significantly increased among the current smoking silicotics with PRISm (adjusted HR=2.44, 95% CI: 1.66-3.59), while the risk of developing AFO decreased among former smokers who had already quitted smoking no matter whether the subjects presented with (adjusted HR=1.97, 95% CI: 1.35-2.87) or without PRISm (adjusted HR=2.44, 95% CI: 1.66-3.59). There was no evidence of a significant interaction between PRISm and smoking status on the risk of all-cause mortality (p for interaction = 0.35) and development of AFO (p for interaction = 0.89).

**Figure 3.**
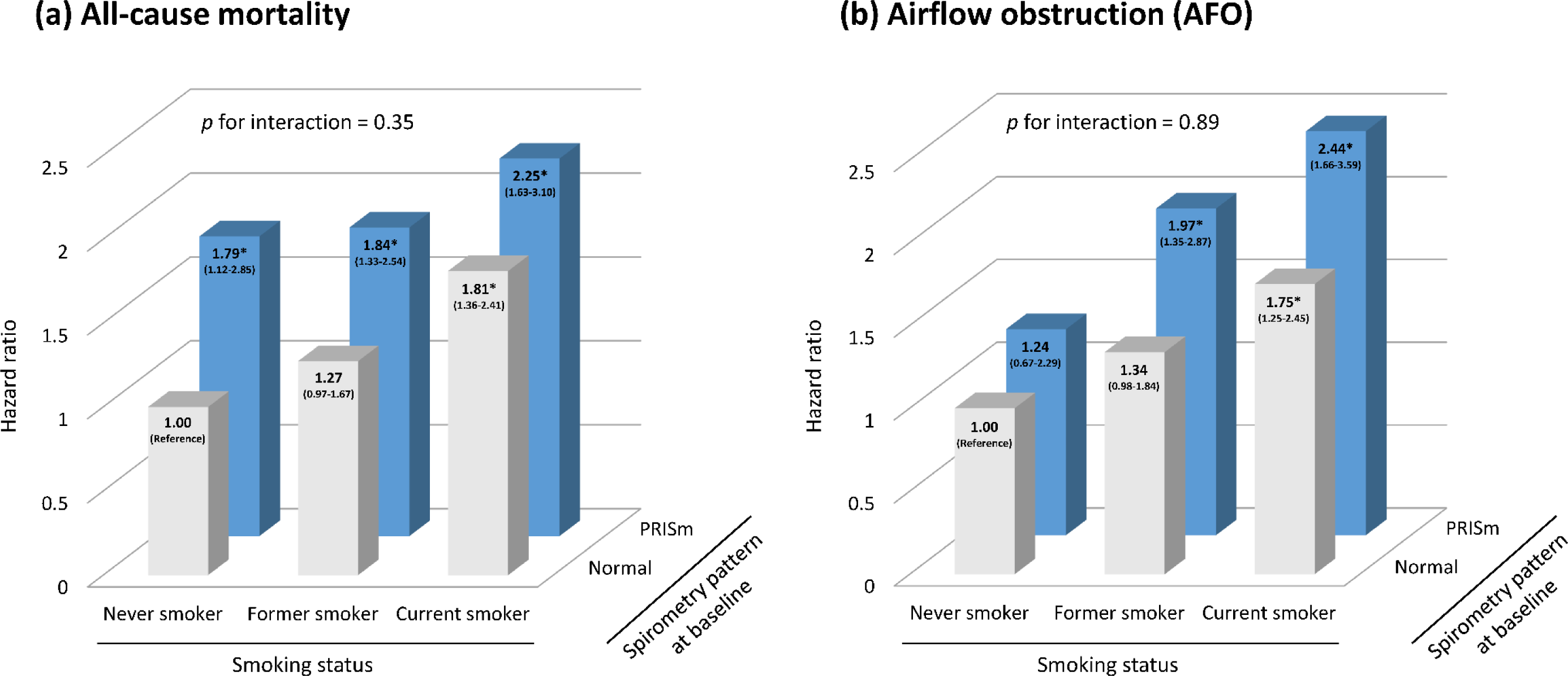
The combined influence of preserved ratio impaired spirometry (PRISm) at baseline and smoking on (a) the risk of all-cause death and (b) the development of airflow obstruction (AFO) Never smokers with normal spirometry at baseline were used as reference. Data are presented as hazard ratio with 95% confidence interval adjusting for age, BMI, history of tuberculosis, cumulative silica exposure, pack-years, and radiographic signs. Abbreviations: AFO, airflow obstruction; PRISm, preserved ratio impaired spirometry; BMI, body mass index.

## Discussion

The present study demonstrated that both PRISm and AFO were significantly associated with increased risk of all-cause death and the mortality from respiratory diseases among the individuals with silicosis. Compared with the silicotic individuals with normal spirometry, those with PRISm had 63% and 74% increased risk of all-cause and respiratory-related mortality, respectively, and significantly greater risk of developing AFO. The magnitude of associations of PRISm with both all-cause death and incident AFO was higher among subjects with a current habit of smoking than those who had quitted smoking or never smoked, although there was no significant interaction between PRISm and smoking status. To our knowledge, this is the first longitudinal cohort study focusing on the risk of mortality in the silicotic population.

While the presence of PRISm has been shown to be a significant predictor of earlier mortality in the general population, the present study focused on the patients with silicosis and confirmed the relationship between PRISm and all-cause as well as respiratory related mortality in this population. In this study, we observed a significantly increased risk of all-cause mortality in the silicotics with PRISm, which was in accordance with the reports of previous general population-based cohort studies in Netherlands^12^, United States^10^, United Kingdom^32^ and Japan^34^, and the magnitude of risk among the silicotics was almost comparable to that in general population^9^. Subgroup analyses by radiographic signs and respiratory symptoms revealed the robustness of this association among the silicotics with or without symptoms and at different stages of silicosis in respect to the lesion progression. Besides, our data indicated that non-restrictive PRISm, which was classified as normal in the conventional criteria of restrictive and obstructive defects, was also associated with significantly increased all-cause mortality but with distinct magnitude of risk from restrictive PRISm, suggesting the different mechanisms of death between restrictive and non-restrictive PRISm^37,38^. Therefore, multidimensional assessment and tailored management of PRISm shall be recommended.

The association between PRISm and cause-specific mortality in the silicotic individuals is not completely consistent with that in the general population. As Wan et al.’s previously reported in a population-based pooled cohort study^10^, our study provided supportive evidence on a relationship between PRISm and mortality from non-malignant respiratory diseases, which are the first leading causes of death in the silicotics^25^. Although the exact mechanism underlying the association between PRISm and respiratory-related mortality has not been fully elucidated, our data supported the early hypotheses that PRISm may be a precursor of COPD^39^. However, despite a significant association of PRISm with cardiovascular events reported by Guerra et al.^20^, Wan et al.^10^ and Wijnant et al.^18^ in the general population, we did not observe a significant association between PRISm and cardiovascular-related mortality among the silicotics, which might be influenced by the “healthy worker” effect because the silicotic workers engaging in tunneling, construction, quarrying or other dust-exposed occupation may exhibit reduced risk of cardiovascular events due to their more physical fitness and better cardiopulmonary function than that of the general population. These findings highlight the clinical importance of identifying individuals with PRISm in the occupational settings as well as in the general population to reduce respiratory-related mortality.

Smoking has been determined as a predictor of mortality and a risk factor of AFO in several independent cohorts^40,41^, which may be partially attributed to the smoking-related bronchial and systemic chronic inflammation that exacerbate the impairment of lung function and finally increase the risk of mortality^42,43^. Our study observed a higher risk of all-cause mortality and incident AFO in current smokers than that in former smokers and never smokers no matter whether the subjects manifested with or without PRISm, and these findings were consistent with those obtained from the Hisayama Study^17^. However, the effect of smoking on all-cause mortality and AFO were independent of PRISm, indicating that smoking may not involve in the progression and prognosis of PRISm. These findings revealed that smoking cessation would be helpful in reducing the risk of all-cause mortality and incident AFO, especially in the occupational population who usually possesses heavier smoking and earlier initiation of smoking than the general population^23,44^.

The strengths of this study include (1) a territory-wide coverage of study population, i.e., individuals with silicosis, (2) a long period of follow-up with a high follow-up rate, and (3) the integrity of data on spirometry, vital status, lifelong smoking habits, and radiographic signs. However, several limitations should be considered. Firstly, lack of postbronchodilator spirometry may lead to the overestimation of some obstructive lung diseases such as COPD. However, this should be of less importance because the focus of this study was on the subjects with a preserved ratio of FEV_1_/FVC instead of COPD. Secondly, the cumulative silica exposure was not directly measured at individual level but estimated using JEM . Nevertheless, the JEM used for this project is a validated tool developed by OSHA and also this tool has been widely used and accepted in occupational epidemiological studies with satisfying power of exposure estimation^45,46^, and hence the potential misclassification of exposure assessment, if it is present in this study, would be nondifferential and lead the risk estimate towards the null. Thirdly, as a distinctive feature of PRISm is the increased rates of transitions to both normal and obstructed spirometry over time^18,47^, analyses was limited to the investigation of associations between cross-sectional (baseline) PRISm and prospective clinical outcomes. Thus, future studies addressing whether distinct longitudinal trajectories within PRISm were differentially associated with clinical outcomes in the silicotic population are warranted.

## Conclusions

The present study demonstrated that PRISm was associated with a significantly increased risk of all-cause mortality, adverse respiratory outcomes, and incident AFO, and this association was not modified by tobacco smoking. These findings suggest the importance of recognizing PRISm in the occupational settings and advocate the continued effort in elucidating the pathophysiological disturbances that involve in the development, progression, and management of PRISm.

## Declaration

## Data Availability

The participants of this study did not give written consent for their data to be shared publicly, so due to the sensitive nature of the research supporting data is not available but it can be available from the corresponding author on reasonable request and with the permission of Hong Kong Department of Health.

## Acknowledgements

The authors are grateful to the Pneumoconiosis Clinic, Tuberculosis and Chest Service, for their support to this study. The authors also appreciate Ocean Mo, Mika Mang and Jade Lee for their assistance in data collection, especially during the hard time of COVID-19 pandemic.

