## Supplementary material for "Association of preserved ratio impaired spirometry with mortality and airflow obstruction in the silicotics: a longitudinal cohort study": Table E1

**Table E1. Inclusion and exclusion of the study subjects**

| Study population | Subjects No. (%) |
|---|---|
| Excluded subjects | 166 (3.7%) |
| > 80 years old# | 83 (1.9%) |
| No record of baseline examination | 28 (0.6%) |
| Invalid spirometry data* | 55 (1.2%) |
| Included subjects | 4315 (96.3%) |
| Total | 4481 (100.0%) |

### Prediction formulae for reference values are applicable for subjects aged 18-80 years, thus those aged over 80 years were excluded[1].

* The spirometry results were considered as invalid when (1) $FEV_1$: < 0.2 L/s or > 7.0 L/s; (2) $FEV_1$ % predicted: < 10% or > 140%; (3) FVC: < 0.2 L or > 7.0 L; (4) FVC % predicted: < 10% or > 140%; or (5) $FEV_1$/FVC ratio: < 0.1 or > 1.0[2]

[1] Ip MS, Ko FW, Lau AC, et al. Updated spirometric reference values for adult Chinese in Hong Kong and implications on clinical utilization. *Chest.* 2006;129(2):384-392. doi:10.1378/chest.129.2.384

[2] Josephs L, Culliford D, Johnson M, Thomas M. Improved outcomes in ex-smokers with COPD: a UK primary care observational cohort study. *Eur Respir J.* 2017;49(5):1602114. Published 2017 May 23. doi:10.1183/13993003.02114-2016
