## Supplementary material for "Association of preserved ratio impaired spirometry with mortality and airflow obstruction in the silicotics: a longitudinal cohort study": Table E2

**Table E2. Hazard ratios (HR) and 95% confidence intervals (CI) for all-cause and respiratory-related mortality by lung function categories (with restrictive/non-restrictive PRISm)**

| Lung function categories | No. of deaths/ subjects | Crude mortality rate* | Crude HR (95% CI) | Adjusted HR (95% CI) | | |
|---|---|---|---|---|---|---|
| | | | Model 0 | Model 1 | Model 2 | Model 3 |
| All-cause mortality (A00-Z99) | | | | | | |
| Normal spirometry | 977/2094 | 30.8 | 1.00 (Ref.) | 1.00 (Ref.) | 1.00 (Ref.) | 1.00 (Ref.) |
| Non-restrictive PRISm | 76/119 | 40.8 | 1.39 (1.10, 1.75) | 1.42 (1.13, 1.80) | 1.40 (1.10, 1.77) | 1.31 (1.03, 1.66) |
| Restrictive PRISm | 273/389 | 57.6 | 2.03 (1.78, 2.32) | 1.92 (1.68, 2.20) | 1.92 (1.68, 2.20) | 1.76 (1.53, 2.02) |
| AFO GOLD 1 | 219/435 | 43.2 | 1.69 (1.46, 1.96) | 1.34 (1.15, 1.55) | 1.30 (1.12, 1.51) | 1.29 (1.11, 1.50) |
| AFO GOLD 2 | 530/825 | 53.9 | 2.08 (1.87, 2.32) | 1.70 (1.52, 1.90) | 1.64 (1.47, 1.84) | 1.57 (1.40, 1.76) |
| AFO GOLD 3 | 245/350 | 80.1 | 3.52 (3.06, 4.06) | 2.62 (2.25, 3.03) | 2.46 (2.11, 2.85) | 2.33 (2.00, 2.72) |
| AFO GOLD 4 | 79/103 | 152.3 | 8.68 (6.87, 10.96) | 6.04 (4.74, 7.70) | 5.95 (4.65, 7.62) | 5.75 (4.45, 7.41) |
| Respiratory-related mortality (J00-J99) | | | | | | |
| Normal spirometry | 477/2094 | 15.0 | 1.00 (Ref.) | 1.00 (Ref.) | 1.00 (Ref.) | 1.00 (Ref.) |
| Non-restrictive PRISm | 51/119 | 27.4 | 1.89 (1.45, 2.47) | 1.80 (1.37, 2.34) | 1.78 (1.36, 2.33) | 1.64 (1.25, 2.15) |
| Restrictive PRISm | 164/389 | 34.6 | 2.12 (1.77, 2.55) | 2.02 (1.68, 2.42) | 2.02 (1.66, 2.42) | 1.78 (1.48, 2.14) |
| AFO GOLD 1 | 113/435 | 22.3 | 1.44 (1.19, 1.75) | 1.36 (1.11, 1.66) | 1.36 (1.11, 1.66) | 1.34 (1.09, 1.64) |
| AFO GOLD 2 | 314/825 | 32.0 | 2.02 (1.76, 2.32) | 1.87 (1.62, 2.16) | 1.85 (1.59, 2.14) | 1.74 (1.49, 2.02) |
| AFO GOLD 3 | 176/350 | 57.5 | 3.72 (3.09, 4.47) | 3.23 (2.66, 3.93) | 3.16 (2.59, 3.85) | 2.90 (2.36, 3.57) |
| AFO GOLD 4 | 64/103 | 123.4 | 8.08 (5.54, 11.79) | 6.57 (4.41, 9.79) | 6.92 (4.63, 10.35) | 6.32 (4.18, 9.59) |

Abbreviations: AFO, airflow obstruction; PRISm, preserved ratio impaired spirometry; HR, hazard ratio; CI, confidence interval.

Model 0: no adjustments.

Model 1: adjusted for age, BMI category, history of tuberculosis, and cumulative silica exposure.

Model 2: adjusted for the covariates in Model 1 plus smoking status and pack-years.

Model 3: adjusted for the covariates in Model 2 plus the radiographic signs, including shape, size, profusion of the small opacities and progressive massive fibrosis.

* Per $10^3$ person-years.
