## Supplementary material for "Association of preserved ratio impaired spirometry with mortality and airflow obstruction in the silicotics: a longitudinal cohort study": Table E3

**Table E3. Subgroup analyses of the association of baseline lung function categories with risk of all-cause mortality by shape of small opacities**

| Lung function category | No. of deaths/ subjects | Crude mortality rate* | Crude HR (95% CI) | Adjusted HR (95% CI) | | |
|---|---|---|---|---|---|---|
| | | | Model 0 | Model 1 | Model 2 | Model 3 |
| *Analysis for subjects with small round opacities (n=2873)* | | | | | | |
| Normal spirometry | 627/1436 | 29.6 | 1.00 (Ref.) | 1.00 (Ref.) | 1.00 (Ref.) | 1.00 (Ref.) |
| PRISm | 229/337 | 50.8 | 1.79 (1.54, 2.08) | 1.73 (1.49, 2.02) | 1.70 (1.46, 1.98) | 1.70 (1.46, 1.98) |
| AFO GOLD 1 | 138/305 | 43.2 | 1.92 (1.60, 2.32) | 1.57 (1.30, 1.90) | 1.57 (1.30, 1.90) | 1.49 (1.23, 1.80) |
| AFO GOLD 2 | 342/565 | 52.4 | 2.11 (1.85, 2.41) | 1.75 (1.53, 2.01) | 1.73 (1.50, 1.98) | 1.67 (1.45, 1.92) |
| AFO GOLD 3 | 135/195 | 75.3 | 3.47 (2.87, 4.20) | 2.79 (2.30, 3.39) | 2.68 (2.20, 3.26) | 2.54 (2.08, 3.09) |
| AFO GOLD 4 | 26/35 | 139.1 | 8.47 (5.66, 12.66) | 6.37 (4.23, 9.58) | 6.13 (4.07, 9.24) | 6.30 (4.16, 9.54) |
| *Analysis for subjects with small irregular opacities (n=1393)* | | | | | | |
| Normal spirometry | 337/639 | 33.02487 | 1.00 (Ref.) | 1.00 (Ref.) | 1.00 (Ref.) | 1.00 (Ref.) |
| PRISm | 116/165 | 56.15829 | 1.87 (1.51, 2.31) | 1.86 (1.50, 2.30) | 1.86 (1.49, 2.28) | 1.83 (1.48, 2.27) |
| AFO GOLD 1 | 78/125 | 43.09749 | 1.43 (1.11, 1.82) | 1.12 (0.87, 1.44) | 1.10 (0.86, 1.42) | 1.09 (0.84, 1.40) |
| AFO GOLD 2 | 184/252 | 57.76681 | 2.05 (1.71, 2.45) | 1.64 (1.36, 1.97) | 1.61 (1.33, 1.94) | 1.55 (1.28, 1.87) |
| AFO GOLD 3 | 105/149 | 90.4346 | 3.64 (2.91, 4.55) | 2.53 (2.01, 3.20) | 2.51 (1.99, 3.17) | 2.40 (1.89, 3.03) |
| AFO GOLD 4 | 49/63 | 158.6068 | 8.09 (5.93, 11.03) | 5.75 (4.14, 7.99) | 5.63 (4.05, 7.84) | 5.12 (3.66, 7.15) |

Abbreviations: AFO, airflow obstruction; PRISm, preserved ratio impaired spirometry; HR, hazard ratio; CI, confidence interval.
Model 0: no adjustments.
Model 1: adjusted for age and BMI category.
Model 2: adjusted for the covariates in Model 1 plus history of tuberculosis, and cumulative silica exposure.
Model 3: adjusted for the covariates in Model 2 plus smoking status and pack-years.
* Per $10^3$ person-years.
