## Supplementary material for "Association of preserved ratio impaired spirometry with mortality and airflow obstruction in the silicotics: a longitudinal cohort study": Table E4

**Table E4. Subgroup analyses of the association of baseline lung function categories with risk of all-cause mortality by profusion of small opacities**

| Lung function category | No. of deaths/ subjects | Crude mortality rate* | Crude HR (95% CI) | Adjusted HR (95% CI) | | |
|---|---|---|---|---|---|---|
| | | | Model 0 | Model 1 | Model 2 | Model 3 |
| *Analysis for subjects with profusion of small opacities in Category 1 (n=2384)* | | | | | | |
| Normal spirometry | 436/1280 | 25.4 | 1.00 (Ref.) | 1.00 (Ref.) | 1.00 (Ref.) | 1.00 (Ref.) |
| PRISm | 81/166 | 39.3 | 1.57 (1.24, 1.99) | 1.47 (1.16, 1.87) | 1.45 (1.14, 1.84) | 1.37 (1.07, 1.74) |
| AFO GOLD 1 | 100/271 | 37.6 | 1.92 (1.54, 2.40) | 1.47 (1.17, 1.84) | 1.46 (1.17, 1.83) | 1.34 (1.07, 1.69) |
| AFO GOLD 2 | 230/439 | 50.7 | 2.45 (2.09, 2.88) | 1.88 (1.59, 2.23) | 1.85 (1.56, 2.19) | 1.76 (1.49, 2.09) |
| AFO GOLD 3 | 101/176 | 74.1 | 3.98 (3.19, 4.97) | 2.66 (2.11, 3.35) | 2.60 (2.06, 3.28) | 2.46 (1.95, 3.11) |
| AFO GOLD 4 | 32/52 | 133.9 | 9.29 (6.42, 13.44) | 5.82 (3.99, 8.48) | 5.70 (3.91, 8.31) | 5.89 (4.02, 8.64) |
| *Analysis for subjects with profusion of small opacities in Category 2 (n=1528)* | | | | | | |
| Normal spirometry | 447/695 | 35.9 | 1.00 (Ref.) | 1.00 (Ref.) | 1.00 (Ref.) | 1.00 (Ref.) |
| PRISm | 177/241 | 51.2 | 1.58 (1.33, 1.88) | 1.58 (1.33, 1.89) | 1.58 (1.32, 1.88) | 1.60 (1.34, 1.91) |
| AFO GOLD 1 | 100/138 | 50.4 | 1.64 (1.32, 2.04) | 1.35 (1.08, 1.68) | 1.34 (1.07, 1.68) | 1.32 (1.06, 1.65) |
| AFO GOLD 2 | 237/305 | 56.7 | 1.89 (1.62, 2.22) | 1.63 (1.38, 1.92) | 1.61 (1.37, 1.90) | 1.59 (1.35, 1.88) |
| AFO GOLD 3 | 99/114 | 97.5 | 4.06 (3.25, 5.07) | 3.29 (2.61, 4.14) | 3.29 (2.61, 4.15) | 3.13 (2.47, 3.96) |
| AFO GOLD 4 | 32/35 | 171.5 | 9.22 (6.4, 13.29) | 7.91 (5.36, 11.69) | 7.66 (5.17, 11.35) | 7.37 (4.95, 10.99) |
| *Analysis for subjects with profusion of small opacities in Category 3 (n=350)* | | | | | | |
| Normal spirometry | 81/100 | 45.95 | 1.00 (Ref.) | 1.00 (Ref.) | 1.00 (Ref.) | 1.00 (Ref.) |
| PRISm | 86/95 | 81.41 | 2.07 (1.52, 2.82) | 2.02 (1.47, 2.78) | 1.94 (1.41, 2.68) | 1.77 (1.27, 2.46) |
| AFO GOLD 1 | 16/20 | 50.01 | 1.15 (0.67, 1.97) | 1.04 (0.60, 1.79) | 1.10 (0.63, 1.90) | 1.06 (0.61, 1.85) |
| AFO GOLD 2 | 59/70 | 63.21 | 1.55 (1.10, 2.18) | 1.42 (1.01, 2.02) | 1.44 (1.01, 2.05) | 1.24 (0.86, 1.78) |
| AFO GOLD 3 | 40/54 | 69.69 | 1.87 (1.27, 2.75) | 1.42 (0.95, 2.14) | 1.34 (0.89, 2.02) | 1.21 (0.79, 1.84) |
| AFO GOLD 4 | 11/11 | 156.41 | 5.32 (2.79, 10.14) | 3.14 (1.58, 6.23) | 3.22 (1.59, 6.52) | 2.96 (1.45, 6.06) |

Abbreviations: AFO, airflow obstruction; PRISm, preserved ratio impaired spirometry; HR, hazard ratio; CI, confidence interval.

Model 0: no adjustments.

Model 1: adjusted for age and BMI category.

Model 2: adjusted for the covariates in Model 1 plus history of tuberculosis, and cumulative silica exposure.

Model 3: adjusted for the covariates in Model 2 plus smoking status and pack-years.

* Per $10^3$ person-years.
