## Supplementary material for "Association of preserved ratio impaired spirometry with mortality and airflow obstruction in the silicotics: a longitudinal cohort study": Table E5

**Table E5. Subgroup analyses of the association of baseline lung function categories with risk of all-cause mortality by size of small opacities**

| Lung function category | No. of deaths/ subjects | Crude mortality rate* | Crude HR (95% CI) | Adjusted HR (95% CI) | | |
|---|---|---|---|---|---|---|
| | | | Model 0 | Model 1 | Model 2 | Model 3 |
| *Analysis for subjects with small opacities in size p or s (n=1796)* | | | | | | |
| Normal spirometry | 403/974 | 29.7 | 1.00 (Ref.) | 1.00 (Ref.) | 1.00 (Ref.) | 1.00 (Ref.) |
| PRISm | 89/162 | 41.4 | 1.51 (1.20, 1.90) | 1.44 (1.14, 1.82) | 1.41 (1.11, 1.78) | 1.39 (1.10, 1.76) |
| AFO GOLD 1 | 93/215 | 39.4 | 1.67 (1.33, 2.09) | 1.23 (0.97, 1.56) | 1.20 (0.94, 1.51) | 1.13 (0.89, 1.44) |
| AFO GOLD 2 | 163/298 | 52.8 | 2.28 (1.89, 2.74) | 1.74 (1.44, 2.11) | 1.71 (1.41, 2.07) | 1.64 (1.36, 1.99) |
| AFO GOLD 3 | 56/110 | 70.5 | 3.40 (2.56, 4.51) | 2.45 (1.84, 3.27) | 2.40 (1.80, 3.20) | 2.31 (1.72, 3.10) |
| AFO GOLD 4 | 21/37 | 155.4 | 13.05 (8.23, 20.69) | 8.92 (5.47, 14.56) | 9.05 (5.54, 14.80) | 9.51 (5.81, 15.56) |
| *Analysis for subjects with small opacities in size q or t (n=2092)* | | | | | | |
| Normal spirometry | 505/1008 | 30.8 | 1.00 (Ref.) | 1.00 (Ref.) | 1.00 (Ref.) | 1.00 (Ref.) |
| PRISm | 195/270 | 53.7 | 1.85 (1.57, 2.18) | 1.85 (1.56, 2.18) | 1.83 (1.55, 2.17) | 1.84 (1.56, 2.18) |
| AFO GOLD 1 | 107/189 | 45.4 | 1.74 (1.41, 2.15) | 1.40 (1.13, 1.74) | 1.40 (1.13, 1.74) | 1.37 (1.10, 1.69) |
| AFO GOLD 2 | 281/406 | 52.9 | 1.95 (1.68, 2.26) | 1.66 (1.43, 1.93) | 1.64 (1.41, 1.91) | 1.60 (1.37, 1.86) |
| AFO GOLD 3 | 135/172 | 80.3 | 3.44 (2.83, 4.17) | 2.65 (2.17, 3.24) | 2.62 (2.14, 3.21) | 2.47 (2.01, 3.03) |
| AFO GOLD 4 | 43/47 | 138.4 | 7.25 (5.29, 9.93) | 5.50 (3.97, 7.61) | 5.37 (3.86, 7.45) | 5.22 (3.74, 7.29) |
| *Analysis for subjects with small opacities in size r or u (n=378)* | | | | | | |
| Normal spirometry | 56/93 | 39.9 | 1.00 (Ref.) | 1.00 (Ref.) | 1.00 (Ref.) | 1.00 (Ref.) |
| PRISm | 61/70 | 77.1 | 1.97 (1.36, 2.85) | 2.02 (1.38, 2.95) | 2.09 (1.42, 3.06) | 2.08 (1.40, 3.09) |
| AFO GOLD 1 | 16/26 | 55.8 | 1.65 (0.94, 2.89) | 1.60 (0.91, 2.81) | 1.69 (0.96, 2.99) | 1.75 (0.98, 3.15) |
| AFO GOLD 2 | 82/113 | 62.6 | 1.76 (1.25, 2.49) | 1.66 (1.18, 2.35) | 1.78 (1.25, 2.53) | 1.83 (1.28, 2.62) |
| AFO GOLD 3 | 49/62 | 102.8 | 3.23 (2.18, 4.80) | 2.64 (1.76, 3.97) | 2.77 (1.84, 4.19) | 2.86 (1.87, 4.37) |
| AFO GOLD 4 | 11/14 | 219.8 | 9.63 (4.92, 18.84) | 7.00 (3.46, 14.16) | 7.97 (3.89, 16.34) | 8.40 (4.00, 17.61) |

Abbreviations: AFO, airflow obstruction; PRISm, preserved ratio impaired spirometry; HR, hazard ratio; CI, confidence interval.

Model 0: no adjustments.

Model 1: adjusted for age and BMI category.

Model 2: adjusted for the covariates in Model 1 plus history of tuberculosis, and cumulative silica exposure.

Model 3: adjusted for the covariates in Model 2 plus smoking status and pack-years.

* Per $10^3$ person-years.
