## Supplementary material for "Association of preserved ratio impaired spirometry with mortality and airflow obstruction in the silicotics: a longitudinal cohort study": Table E6

Model 3: adjusted for the covariates in Model 2 plus smoking status and pack-years.

* Per $10^3$ person-years.

**Table E6. Subgroup analyses of the association of baseline lung function categories with risk of all-cause mortality by progressive massive fibrosis**

| Lung function category | No. of deaths/ subjects | Crude mortality rate* | Crude HR (95% CI) | Adjusted HR (95% CI) | | |
|---|---|---|---|---|---|---|
| | | | Model 0 | Model 1 | Model 2 | Model 3 |
| *Analysis for subjects with small opacities only (without PMF) (n=3516)* | | | | | | |
| Normal spirometry | 815/1832 | 29.4 | 1.00 (Ref.) | 1.00 (Ref.) | 1.00 (Ref.) | 1.00 (Ref.) |
| PRISm | 246/373 | 47.7 | 1.69 (1.47, 1.95) | 1.66 (1.44, 1.92) | 1.64 (1.42, 1.89) | 1.64 (1.42, 1.89) |
| AFO GOLD 1 | 180/360 | 43.0 | 1.75 (1.49, 2.06) | 1.38 (1.17, 1.62) | 1.36 (1.15, 1.61) | 1.32 (1.12, 1.56) |
| AFO GOLD 2 | 383/624 | 52.0 | 2.09 (1.85, 2.37) | 1.75 (1.55, 1.99) | 1.73 (1.52, 1.96) | 1.67 (1.47, 1.89) |
| AFO GOLD 3 | 168/254 | 75.7 | 3.45 (2.92, 4.09) | 2.59 (2.18, 3.08) | 2.54 (2.13, 3.02) | 2.37 (1.99, 2.83) |
| AFO GOLD 4 | 54/73 | 143.7 | 8.54 (6.46, 11.28) | 6.03 (4.51, 8.07) | 5.78 (4.32, 7.74) | 5.66 (4.21, 7.61) |
| *Analysis for subjects with large opacity (with PMF) (n=763)* | | | | | | |
| Normal spirometry | 151/245 | 40.5 | 1.00 (Ref.) | 1.00 (Ref.) | 1.00 (Ref.) | 1.00 (Ref.) |
| PRISm | 100/131 | 69.6 | 1.97 (1.52, 2.54) | 1.93 (1.49, 2.50) | 1.94 (1.50, 2.51) | 1.93 (1.49, 2.51) |
| AFO GOLD 1 | 39/73 | 45.5 | 1.40 (0.98, 2.00) | 1.17 (0.82, 1.68) | 1.17 (0.81, 1.68) | 1.18 (0.82, 1.70) |
| AFO GOLD 2 | 143/194 | 60.7 | 1.75 (1.38, 2.20) | 1.47 (1.16, 1.88) | 1.47 (1.15, 1.87) | 1.48 (1.16, 1.89) |
| AFO GOLD 3 | 75/93 | 94.6 | 3.14 (2.36, 4.18) | 2.59 (1.93, 3.48) | 2.63 (1.95, 3.54) | 2.56 (1.90, 3.45) |
| AFO GOLD 4 | 23/27 | 171.6 | 8.43 (5.31, 13.37) | 7.31 (4.54, 11.76) | 7.37 (4.57, 11.86) | 7.29 (4.51, 11.80) |

Abbreviations: AFO, airflow obstruction; PRISm, preserved ratio impaired spirometry; PMF, progressive massive fibrosis; HR, hazard ratio; CI, confidence interval.

Model 0: no adjustments.

Model 1: adjusted for age and BMI category.

Model 2: adjusted for the covariates in Model 1 plus history of tuberculosis, and cumulative silica exposure.

Model 3: adjusted for the covariates in Model 2 plus smoking status and pack-years.

* Per $10^3$ person-years.

**Table E7. Hazard ratios (HR) and 95% confidence intervals (CI) for all-cause and cause-specific mortality by LLN-defined spirometric lung function class**

| Lung function category | No. of deaths/ subjects | Crude mortality rate* | Crude HR (95% CI) | Adjusted HR (95% CI) | | |
|---|---|---|---|---|---|---|
| | | | Model 0 | Model 1 | Model 2 | Model 3 |
| All-cause mortality | | | | | | |
| Normal spirometry - LLN | 989/2128 | 32.1 | 1.00 (Ref.) | 1.00 (Ref.) | 1.00 (Ref.) | 1.00 (Ref.) |
| PRISm - LLN | 384/581 | 55.9 | 1.91 (1.70, 2.15) | 1.65 (1.45, 1.85) | 1.64 (1.40, 1.82) | 1.51 (1.34, 1.71) |
| AFO - LLN | 1026/1606 | 53.6 | 1.83 (1.68, 2.00) | 1.70 (1.51, 1.81) | 1.65 (1.47, 1.75) | 1.58 (1.45, 1.73) |
| Respiratory-related mortality | | | | | | |
| Normal spirometry - LLN | 472/2128 | 15.3 | 1.00 (Ref.) | 1.00 (Ref.) | 1.00 (Ref.) | 1.00 (Ref.) |
| PRISm - LLN | 226/581 | 32.9 | 2.01 (1.71, 2.36) | 1.80 (1.53, 2.12) | 1.79 (1.52, 2.10) | 1.59 (1.35, 1.88) |
| AFO - LLN | 661/1606 | 34.5 | 2.23 (1.99, 2.50) | 2.03 (1.81, 2.29) | 2.01 (1.79, 2.27) | 1.88 (1.66, 2.12) |
| Lung cancer mortality | | | | | | |
| Normal spirometry - LLN | 134/2128 | 4.4 | 1.00 (Ref.) | 1.00 (Ref.) | 1.00 (Ref.) | 1.00 (Ref.) |
| PRISm - LLN | 32/581 | 4.7 | 0.83 (0.56, 1.22) | 0.79 (0.53, 1.16) | 0.81 (0.55, 1.21) | 0.84 (0.56, 1.26) |
| AFO - LLN | 83/1606 | 4.3 | 0.79 (0.60, 1.04) | 0.77 (0.58, 1.03) | 0.72 (0.54, 0.97) | 0.74 (0.55, 1.00) |
| Cardiovascular-related mortality | | | | | | |
| Normal spirometry - LLN | 82/2128 | 2.7 | 1.00 (Ref.) | 1.00 (Ref.) | 1.00 (Ref.) | 1.00 (Ref.) |
| PRISm - LLN | 32/581 | 4.7 | 1.37 (0.91, 2.05) | 1.30 (0.85, 1.99) | 1.29 (0.84, 1.98) | 1.40 (0.92, 2.16) |
| AFO - LLN | 54/1606 | 2.8 | 0.84 (0.60, 1.19) | 0.88 (0.62, 1.25) | 0.88 (0.62, 1.26) | 0.89 (0.62, 1.29) |

Abbreviations: AFO, airflow obstruction; PRISm, preserved ratio impaired spirometry; HR, hazard ratio; CI, confidence interval.
Model 0: no adjustments.
Model 1: adjusted for age, BMI category, history of tuberculosis, and cumulative silica exposure.
Model 2: adjusted for the covariates in Model 1 plus smoking status and pack-years.
Model 3: adjusted for the covariates in Model 2 plus the radiographic signs, including shape, size, profusion of the small opacities and progressive massive fibrosis.
* Per $10^3$ person-years.
